# The effect of digital-enabled multidisciplinary therapy conferences on efficiency and quality of the decision making in prostate-cancer care

**DOI:** 10.1101/2022.02.23.21268241

**Authors:** Erik Rönmark, Ralf Hoffmann, Viktor Skokic, Maud de Klerk-Starmans, Fredrik Jäderling, Pieter Vos, Maudy C.W. Gayet, Hans Hofstraat, Marco Janssen, Olof Akre, Per H Vincent

## Abstract

**Objectives:** To investigate the impact on efficiency and quality of pre-prostatectomy multidisciplinary therapy conferences (MDT) at Karolinska University Hospital related to the use of a digital solution compared to standard of care. Further, to explore whether gains in MDT efficiency and quality impact oncological or functional patient outcomes.

**Methods:** We conducted a prospective, observational study of preoperative prostate-cancer MDT at Karolinska between February 2017 and March 2021. We compared efficiency and quality of the standard MDT and the MDT using the digital solution *IntelliSpace Precision Medicine Multidisciplinary Team Orchestrator* (ISPM) based on the previously used MDT-MODe approach. Clinical and patient-reported functional outcomes were derived from the medical records and the Swedish National Prostate Cancer Register (NPCR).

**Results:** While ISPM was used during the multidisciplinary therapy conference, the time spent per patient was reduced by 24 % (p<0.001) and most of the MDT-MODe items were scored significantly higher. There was a reduction in pelvic lymph-node dissection procedures in the ISPM cohort (p=0.001) and an increased proportion of unilateral nerve-sparing procedures (p=0.005), while all other outcome-related measures were not significantly different between the two patient groups.

**Discussion & Conclusion:** To increase the value of the MDT, all data relevant for treatment decision need to be purposefully presented and compiled, which also enables secondary use of the data.

The use of a digital solution during preoperative MDTs for prostate-cancer decision-making at Karolinska University Hospital improved the efficiency and quality of this multidisciplinary team meeting without impacting patient outcomes.

**What is already known?:**

- Multidisciplinary therapy conferences are widely used in modern cancer care.
- Patients discussed in a multidisciplinary therapy conference are more likely to receive appropriate staging and treatment.
- The multidisciplinary therapy conference is time consuming and rarely digitalized or adequately structured.

**What does this paper add?:**

- The use of a digital clinical decision support system during preoperative prostate-cancer multidisciplinary therapy conferences improved the efficiency and quality of the meetings.
- The introduction of this digital clinical decision support system was not associated with changes in oncological and functional outcomes after surgery.

## INTRODUCTION

The multidisciplinary therapy conference (MDT) has become a corner stone of cancer care. Patients who are discussed in an MDT, where a team of specialist physicians and nurses gather to summarize relevant data and decide on treatment recommendations, are more likely to receive appropriate staging and treatment plans, but it is unclear whether this also results in improved patient outcomes.^1^ A recent study has indicated that access to complete case information and clarified roles of healthcare professionals are vital prerequisites for a systematic MDT approach.^2^

The MDT often gathers a large number of health professionals, and, with more complex diagnostic and therapeutic options, the quality and efficiency of the decision-making process becomes increasingly important. MDT conferences are rarely fully digitalized or adequately structured, which may affect the quality and efficiency of the decision-making process. Data are not compiled and presented visually in a structured way and clinical parameters are presented verbally, which may lead to delays in the discussion when information needs to be repeated. Lack of continuous access to the clinical parameters during the MDT session may lead to information loss and hamper the multidisciplinary character of the MDT, thereby increasing the risk of non-optimal treatment decisions. Moreover, if the consensus decisions are not captured in the electronic medical records (EMR) in real-time, this may lead to errors, misunderstandings, and delay in data transfer to the EMR.

With the field of digital health evolving rapidly, solutions for MDTs have been developed and assessed. Structuring MDTs by use of such solutions has been shown to increase adherence to national guidelines and efficiency in several tumour forms.^3 4^

To increase the MDT efficiency without compromising quality of patient care, multiple quality-assessment tools and discussion checklists have been developed. Whether these tools also positively impact patient outcomes remains unknown.^5^ In this study, we hypothesized that use of a digital, patient-centric, diagnosis-specific solution developed jointly by us (*IntelliSpace Precision Medicine Multidisciplinary Team Orchestrator*, further referred to as ‘ISPM’ throughout this text) during pre-prostatectomy MDTs at Karolinska University Hospital would improve the efficiency and quality of the MDT. The primary aim of the study was to investigate whether the use of the ISPM application saved meeting time and improved the quality of the decision process. The secondary aim was to assess whether the oncological and functional patient outcomes were affected by the implementation of ISPM.

## METHODS

We have done a prospective observational cohort study comparing patient cohorts before and after the introduction of the clinical decision-support tool ISPM. The study was conducted between February 2017 and March 2021 at Karolinska University Hospital including patients discussed at pre-prostatectomy MDTs before undergoing robot-assisted radical prostatectomy.

### Study setting

Hospital care in Sweden is entirely funded by taxes, and is therefore, as a rule, population based. Karolinska University Hospital is a Swedish tertiary referral hospital treating patients in all risk categories but with emphasis on high-risk patients referred from all regions of Sweden.

The weekly pre-prostatectomy MDT meeting is attended by 10 to 12 specialists in urology and radiology and aims to find a surgical strategy for an optimal balance between radical removal of the prostate cancer and postoperative functional outcomes.

Before we introduced ISPM, a staff urologist, reading from EMR excerpts, verbally reported the clinical data, followed by a presentation of the MR images by a radiologist. The staff then discussed the optimal strategy for degree of nerve-sparing surgery, extent of sphincter sparing dissection in the apex, lymph-node dissection or not, degree of radicality in the bladder neck and the seminal vesicles. The concluded surgical treatment strategy plan was documented by the staff urologist in the EMR after the conference.

After the introduction of ISPM, all relevant clinical and radiological data were entered in the ISPM platform prior to the MDT meeting. In contrast to the baseline setting, clinical and radiological data were continuously visualized on the ISPM dashboard during the MDT meeting alongside the MR images until the surgery treatment plan had been captured in ISPM using the treatment plan documentation tool of the application (Figure 1).

**Figure 1:**
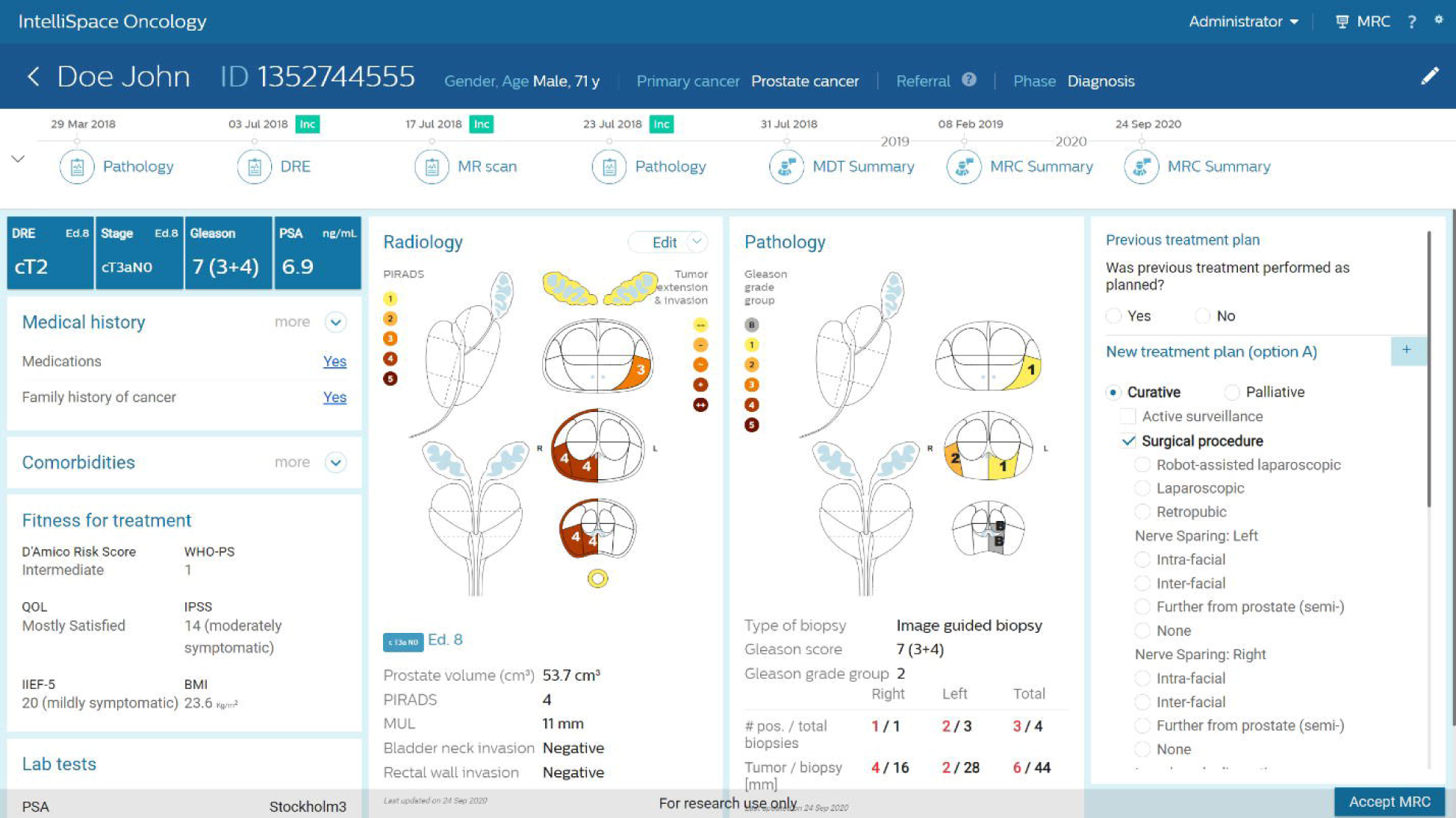
ISPM dashboard as implemented and used in the prospective, observational study on the impact of a digital solution during the MDTs in the prostate-cancer care flow at Karolinska University Hospital. Patient data are fictional and do not originate from a real person.

A baseline measurement in the standard MDT setting (before the use of ISPM) was carried out (Feb 2017-Sept 2019), and, consecutively, data were collected while ISPM was in use (Oct 2019-Mar 2021). The efficiency and quality of the MDTs was compared by timing the discussion and using a modified version of the Metric Decision-Making (MDT-MODe).^6^ Nine items measuring quality were scored using a Likert scale (1, 3 and 5) with higher score indicating higher quality (for details of the modified version of the MDT-MODe used in this study see Supplementary Table 1). We grouped the MDT-MODe items into two main categories: MDT-MODe items relating to the availability and presentation of decision-relevant data, and MDT-MODe items related to the efficiency of MDT execution and team member interaction. Two observers, not participating in the therapy discussion, took turns assigning the MDT-MODe scores. An inter-rater variability analysis was conducted by letting the two observers assign scores to the same MDTs on three separate occasions to ensure agreement.

### Software platform

The *IntelliSpace Precision Medicine Multidisciplinary Team Orchestrator*, or ‘ISPM’, software solution enables preparing, scheduling, visualization, presentation and documentation of information and decisions taken in MDT case discussions. Using SQL queries, the system collects and transforms structured and unstructured data from the hospital data lake into a prostate data model and stores the result into a FHIR database following SNOMED-CT codes. In the study implementation, variables of interest but not available in the research copy of the Karolinska data lake were manually entered in ISPM.

### Patient population

In all, 924 patients were discussed at MDTs in the period Feb 2017-Sept 2019, before the implementation of the ISPM software (“baseline” cohort), and 405 at conferences between Oct 2019 and Mar 2021 using ISPM (“ISPM” cohort). Only patients undergoing prostatectomy as primary treatment for prostate cancer at Karolinska University Hospital within 30 days after their preoperative conference were included, to increase the likelihood that the conference decision was implemented. We assigned MDT-MODe scores to 164 baseline and 163 ISPM patients, at 21 and 22 MDTs, respectively.

The study was approved by the Swedish Ethical Review Authority.

### Oncological and functional outcomes

All clinical and patient-reported outcome data were obtained from routinely collected clinical or quality follow-up data. Positive surgical margin (as sign of remaining cancer and hence non-radical treatment) was used as a surrogate for oncological quality with significant positive margin defined as at least of three millimeters length. Other relevant post- and perisurgical outcomes, such as extended lymph-node dissection, positive lymph nodes, and nerve-sparing surgery, were also analyzed. Functional outcomes were obtained using the questionnaires in the Swedish National Prostate Cancer Register (NPCR) that all prostate-cancer patients in Sweden are invited to answer before undergoing primary treatment and twelve months after treatment. The questionnaires are administered in collaboration with the Swedish Regional Cancer Centers and NPCR and can be found at https://npcr.se/eprom/dokument. In this study, we defined urinary continence as “use of less than one protective urinary pad per day” and urinary incontinence as “use of one or more protective urinary pad per day”. Erectile function was measured using the International Index of Erectile Function questionnaire (IIEF-5)^7^ with erectile dysfunction defined as less than 12 points. Quality of life regarding ‘erectile function satisfaction and continence satisfaction’ was defined as a self-report of either not bothering the patient at all or only to a small degree. Tumour grade was scored using ISUP grading.^8^

### Statistical Analysis

In tables 1-3, comparisons of the characteristics of the studied population with respect to the use of ISPM at MDTs were structured according to the following: the distributions of numerical variables or ordinal variables with more than two levels were compared using the Mann-Whitney U test. The distributions of categorical variables with more than two categories were compared using the χ^2^ test, whereas the distributions of categorical variables with two categories where one category was identified as the outcome of interest were compared in terms of prevalence ratios and the likelihood ratio test associated with an estimated log-binomial model. Levene’s test, centered at the median, was used to assess the difference in variance between non-normally distributed variables.

**Table 1:**
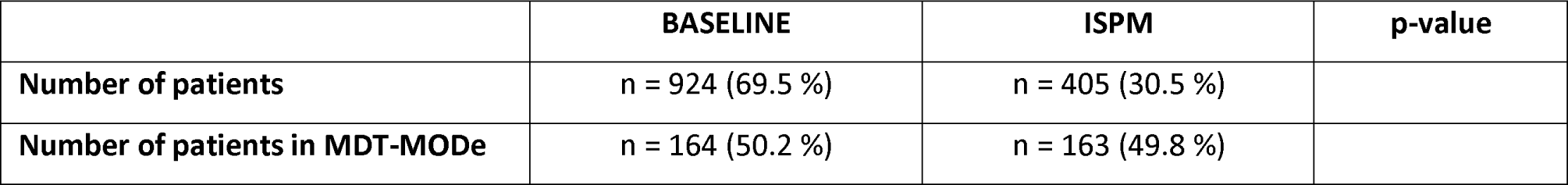
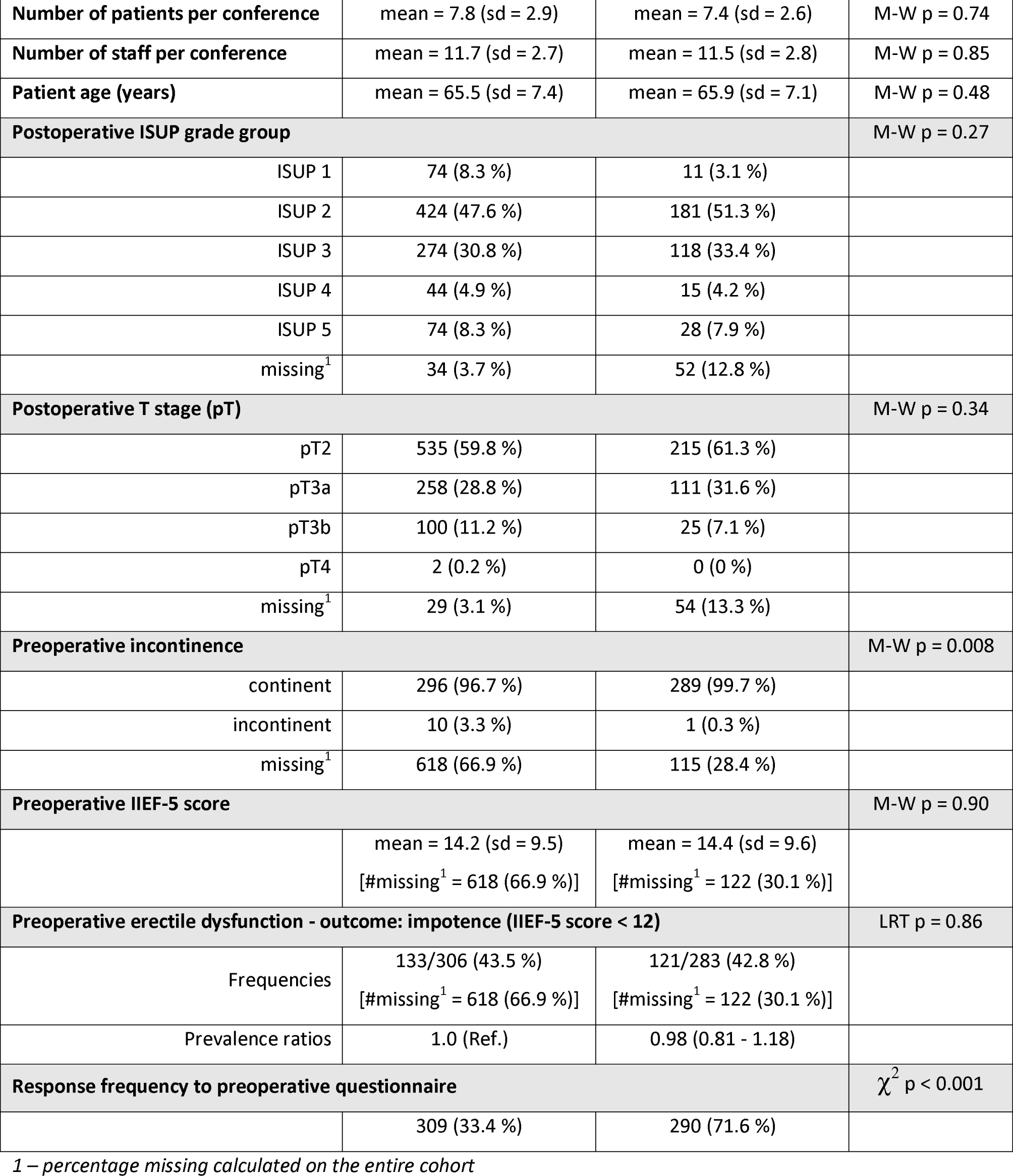
patient demographics of the baseline versus the ISPM cohort

|  | **BASELINE** | **ISPM** | **p-value** |
| --- | --- | --- | --- |
| **Number of patients** | n = 924 (69.5 %) | n = 405 (30.5 %) |  |
| **Number of patients in MDT-MODE** | n = 164 (50.2 %) | n = 163 (49.8 %) |  |
| Number of patients per conference | mean = 7.8 (sd = 2.9) | mean = 7.4 (sd = 2.6) | M-W p = 0.74 |
| Number of staff per conference | mean = 11.7 (sd = 2.7) | mean = 11.5 (sd = 2.8) | M-W p = 0.85 |
| Patient age (years) | mean = 65.5 (sd = 7.4) | mean = 65.9 (sd = 7.1) | M-W p = 0.48 |
| Postoperative ISUP grade group |  |  | M-W p = 0.27 |
| ISUP 1 | 74 (8.3 %) | 11 (3.1 %) |  |
| ISUP 2 | 424 (47.6 %) | 181 (51.3 %) |  |
| ISUP 3 | 274 (30.8 %) | 118 (33.4 %) |  |
| ISUP 4 | 44 (4.9 %) | 15 (4.2 %) |  |
| ISUP 5 | 74 (8.3 %) | 28 (7.9 %) |  |
| missing<sup>1</sup> | 34 (3.7 %) | 52 (12.8 %) |  |
| Postoperative T stage (pT) |  |  | M-W p = 0.34 |
| pT2 | 535 (59.8 %) | 215 (61.3 %) |  |
| pT3a | 258 (28.8 %) | 111 (31.6 %) |  |
| pT3b | 100 (11.2 %) | 25 (7.1 %) |  |
| pT4 | 2 (0.2 %) | 0 (0 %) |  |
| missing<sup>1</sup> | 29 (3.1 %) | 54 (13.3 %) |  |
| Preoperative incontinence |  |  | M-W p = 0.008 |
| continent | 296 (96.7 %) | 289 (99.7 %) |  |
| incontinent | 10 (3.3 %) | 1 (0.3 %) |  |
| missing<sup>1</sup> | 618 (66.9 %) | 115 (28.4 %) |  |
| Preoperative IIEF-5 score |  |  | M-W p = 0.90 |
|  | mean = 14.2 (sd = 9.5)<br>[#missing<sup>1</sup> = 618 (66.9 %)] | mean = 14.4 (sd = 9.6)<br>[#missing<sup>1</sup> = 122 (30.1 %)] |  |
| Preoperative erectile dysfunction - outcome: impotence (IIEF-5 score < 12) |  |  | LRT p = 0.86 |
| Frequencies | 133/306 (43.5 %)<br>[#missing<sup>1</sup> = 618 (66.9 %)] | 121/283 (42.8 %)<br>[#missing<sup>1</sup> = 122 (30.1 %)] |  |
| Prevalence ratios | 1.0 (Ref.) | 0.98 (0.81 - 1.18) |  |
| Response frequency to preoperative questionnaire | | | $\chi^2$ p < 0.001 |
|  | 309 (33.4 %) | 290 (71.6 %) |  |

1 – percentage missing calculated on the entire cohort

For figures 2A and 2C-2F, the distributions of ordinal variables were compared using the Mann-Whitney U test. For figure 2B, the association between the usage of ISPM and the duration of discussion for each individual patient at the MDTs was studied using a linear regression model including the number of patients evaluated at a conference, the usage of ISPM and their interaction as explanatory variables.

**Figure 2A:**
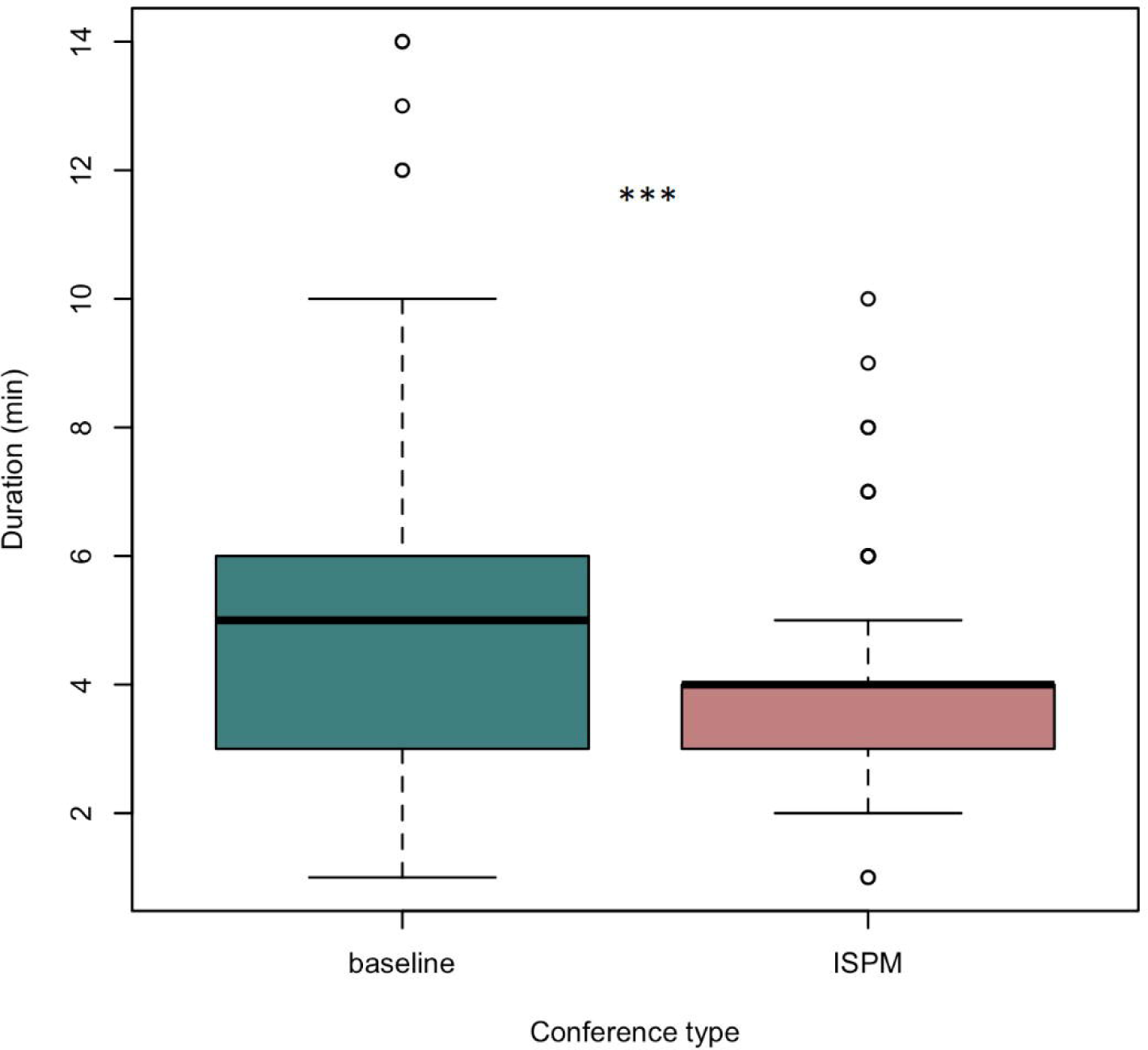
time spent in the MDT meeting per patient in the baseline setting (164 patients) vs. the ISPM setting (163 patients). Box plot with median and IQR; whiskers denote ±1.5 ×IQR. ***=p<0.001

**Figure 2B:**
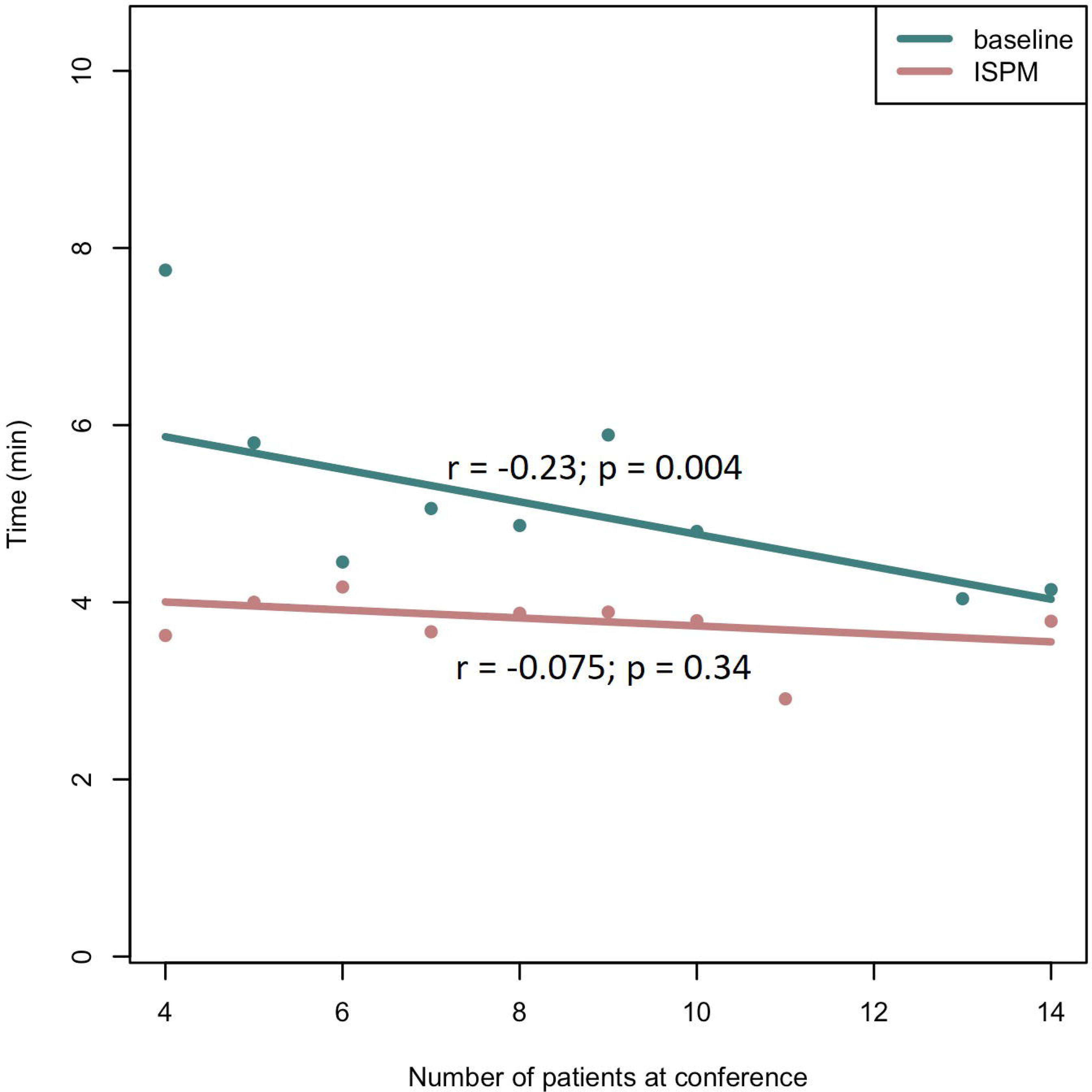
interaction between mean time (minutes) spent per patient and number of patients scheduled and discussed during the MDT. Dots indicate the mean durations at conferences with a particular number of patients being discussed. Regression lines are derived from 164 (baseline) and 163 (ISPM) patients per group.

**Figure 2C:**
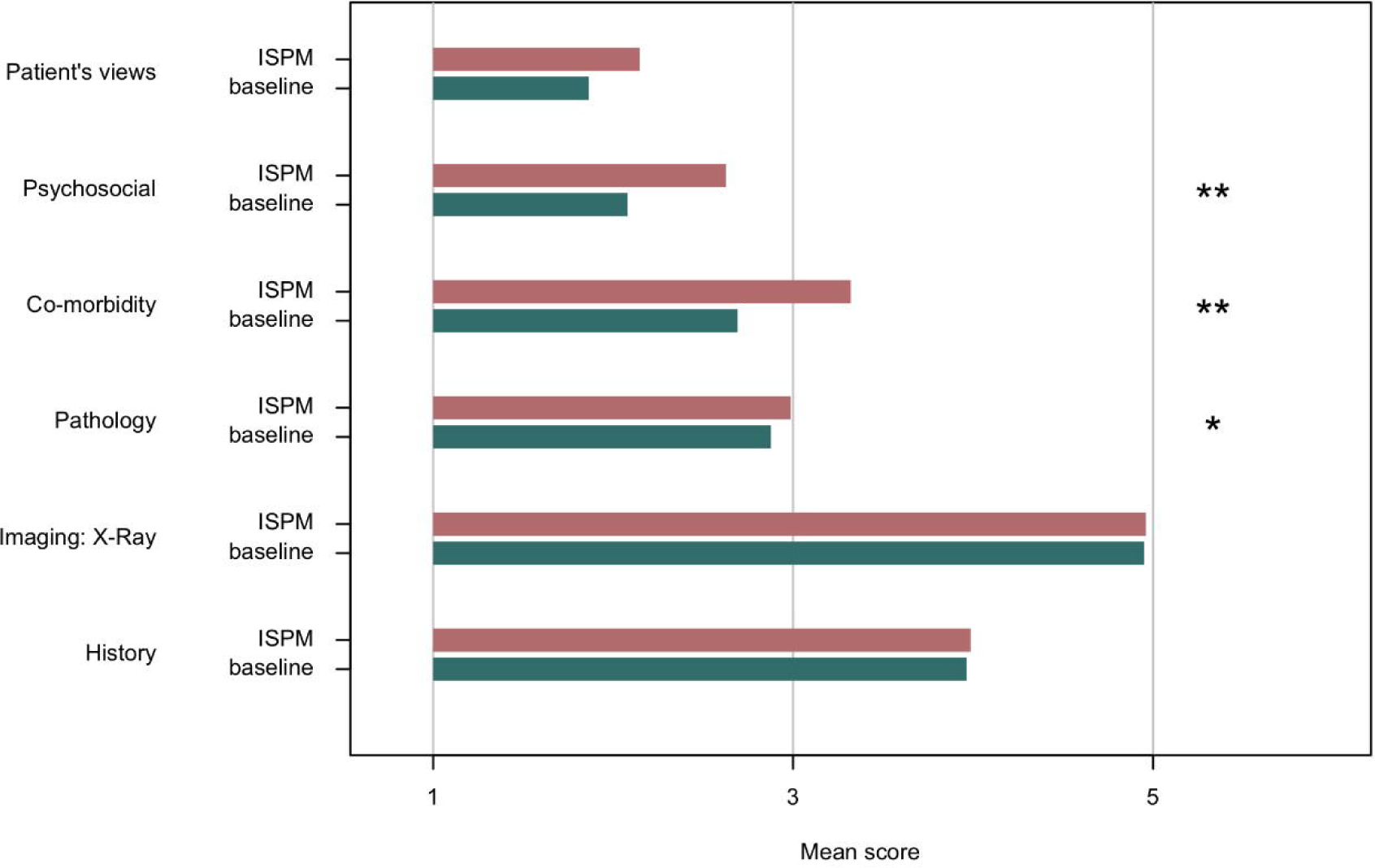
MDT-MODe items concerning information presentation. **=p<0.01; *=p<0.05.

**Figure 2D:**
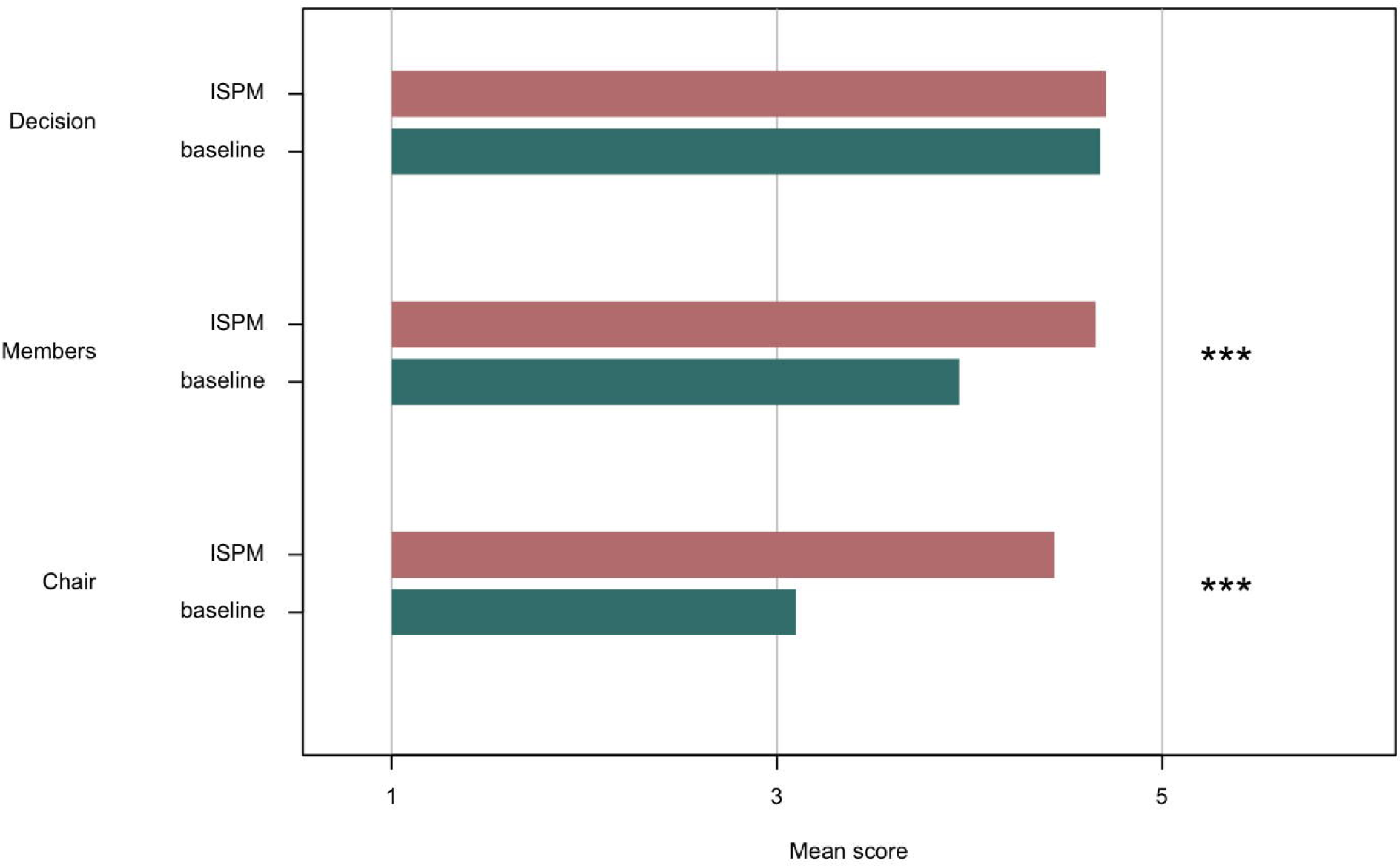
MDT-MODe items concerning leadership and team interaction. ***=p<0.001.

**Figure 2E:**
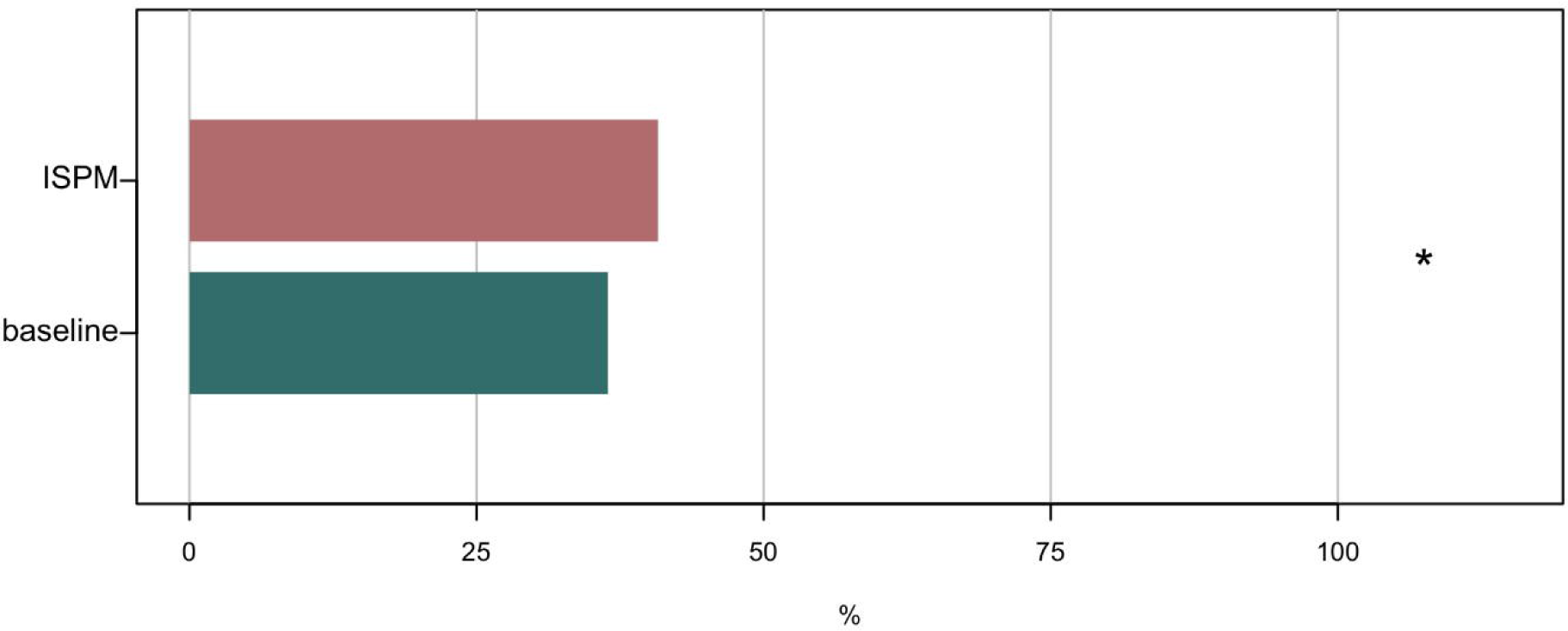
percentage of staff members actively participating per patient case discussion and decision making in the MDT. *=p<0.05

**Figure 2F:**
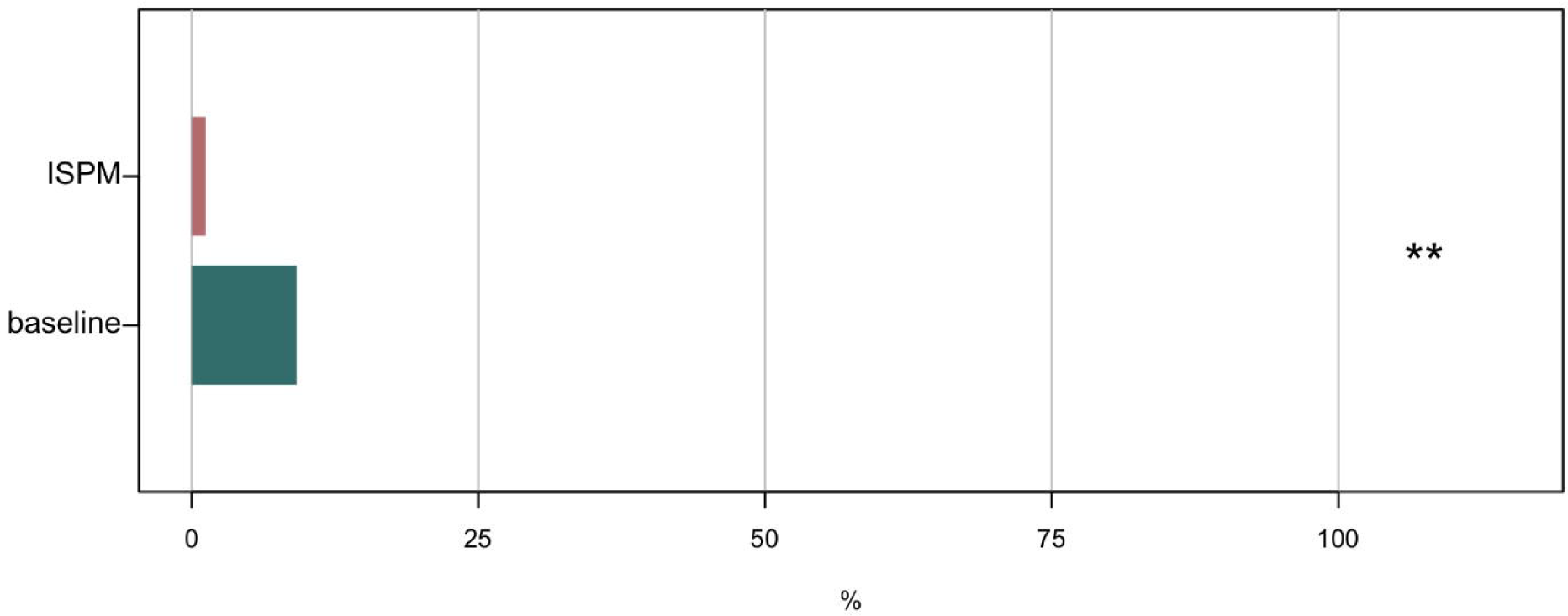
percentage of patients for which questions were raised during the MDT meeting to repeat already presented information. **=p<0.01

All calculations were performed in R version 4.0.0.

## RESULTS

### Patient cohorts

The two cohorts (baseline and ISPM) were similar with respect to demographic and clinical characteristics, mean patient age, postoperative ISUP grade group, tumour stage, and erectile function (Table 1). The response rate to the NPCR questionnaires measuring preoperative functional status was significantly higher among patients discussed using the ISPM solution, 70.9 % versus 32.6 % when ISPM was not used (p<0.001). Among those who responded, incontinence was more common in the baseline group, but the difference was small in absolute numbers, 3.4 % (10 out of 298 patients) compared to 0.3 % (1 out of 287 patients).

### Time efficiency of the MDT

The average time spent discussing each patient was 24 % shorter in the ISPM compared with the baseline setting (3.8 versus 5.0 min; p<0.001; Figure 2A). There was also a significant difference in variances between comparison groups (Levene’s test; p<0.001), indicating a more predictive duration per patient when using ISPM. During the baseline period, the time spent discussing each patient decreased with increasing number of cases in the meeting (Pearson correlation=-0.23; p=0.04). During the ISPM period, there was no such significant correlation (Pearson correlation=-0.075; p=0.36; Figure 2B).

### Quality of the MDT

There were higher MDT-MODe scores for the information presentation items psychosocial, co-morbidity (p<0.01) and pathology (p<0.05) during the ISPM period, while there was no difference in the presentation of patient’s views, imaging, and patient history (Figure 2C). Team interaction items regarding quality of leadership (Chair) and contribution of specialty (Members) also received higher scores in the ISPM setting (p<0.001; Figure 2D). Furthermore, the fraction of participants actively taking part in the MDT discussion increased using ISPM (p<0.05; Figure 2E). Moreover, we observed that there were significantly fewer questions on already presented data raised in the meeting while ISPM was in use (p<0.01; Figure 2F).

### Oncological and functional outcomes

There was no statistically significant difference with respect to oncological outcomes between the baseline and the ISPM cohorts. The proportion of men with positive surgical margins was 27.7 % in the baseline group and 25.9 % in the ISPM group (p=0.66; Table 2; Supplementary Table 2).

**Table 2:**
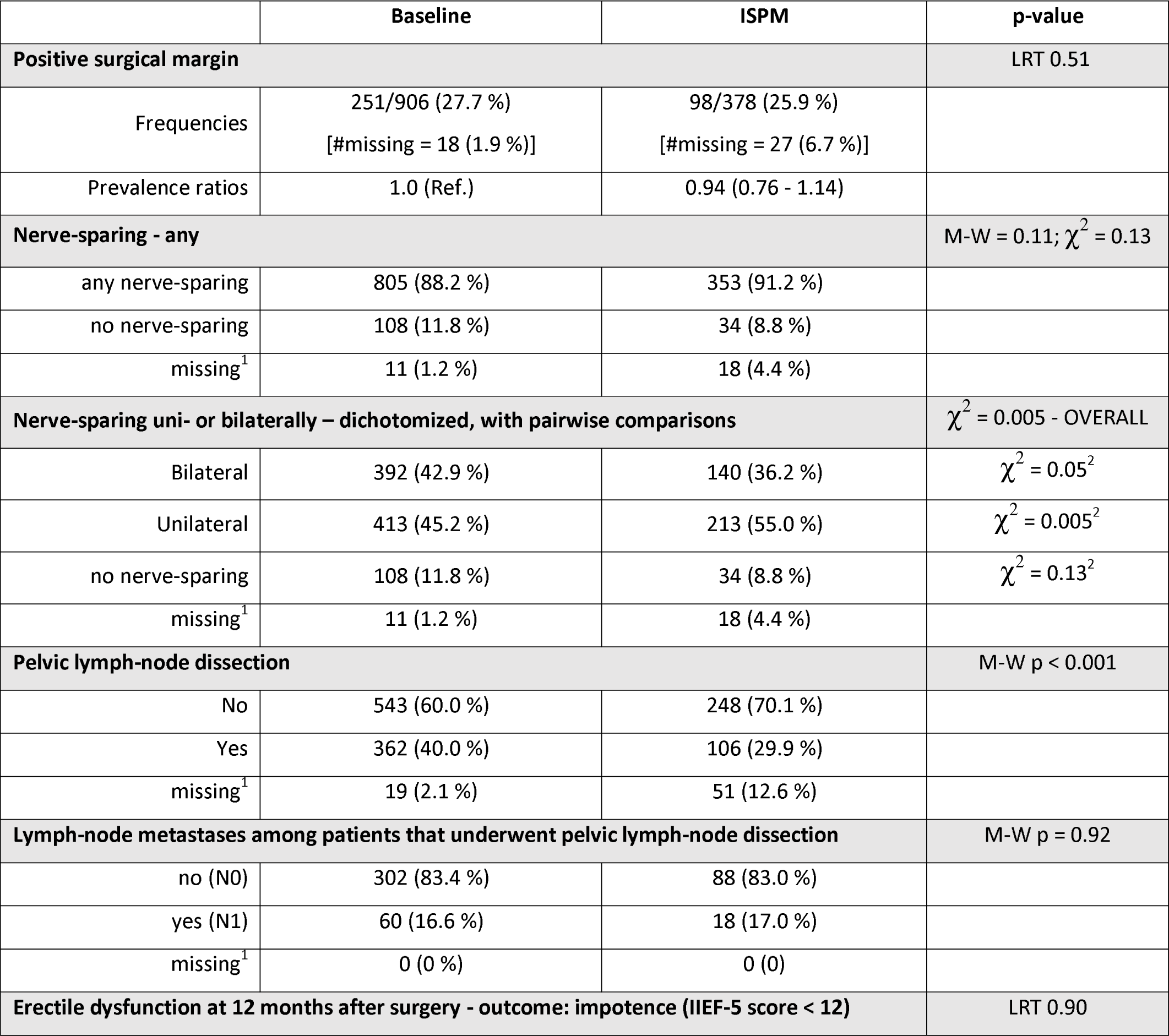
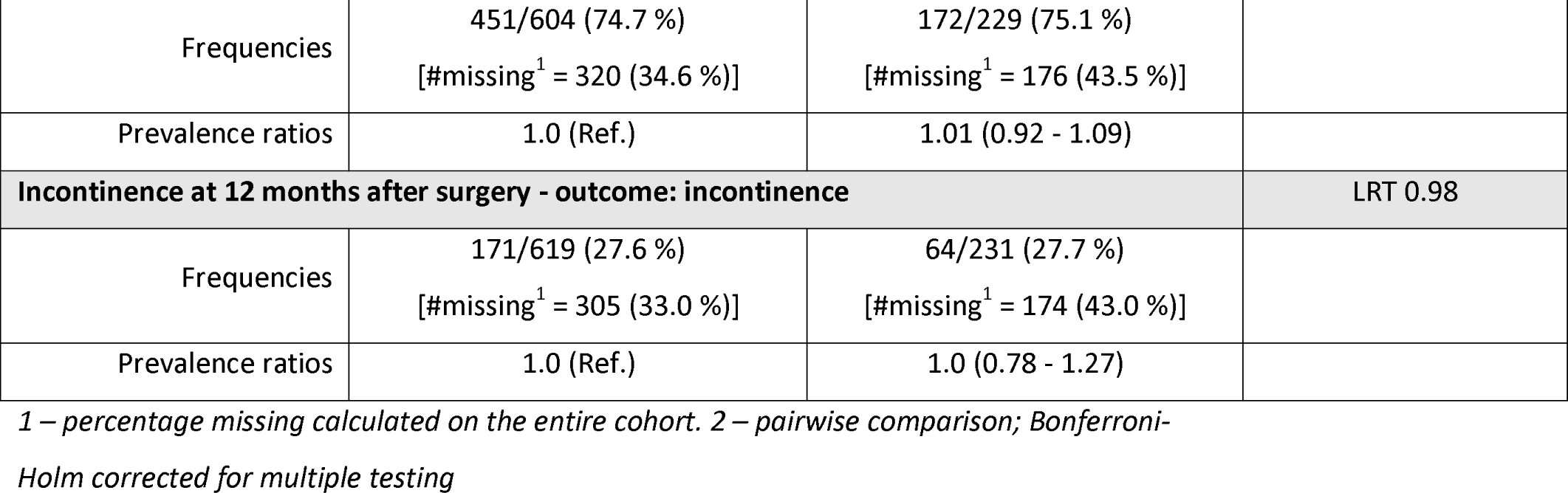
Oncological, perioperative, and 12-month functional (urinary, sexual) patient outcomes of the baseline vs. the ISPM patient cohort

The overall frequency of nerve-sparing and non-nerve-sparing surgery was virtually similar in the comparison groups, but the patterns differed slightly with more unilateral nerve-sparing surgery in the ISPM group and more bilateral nerve-sparing and non-nerve-sparing in the baseline group (Table 2).

An extended pelvic lymph-node dissection was carried out more often in the baseline cohort, 39.1 % vs 26.4 % for the ISPM cohort, respectively (p=0.001; Table 2), but there was no difference in proportion of histologically confirmed metastases (16.6 % in the baseline vs 17.0 % in the ISPM cohort; p=0.92; Table 2).

The functional outcomes one year after surgery were similar for the two groups. Erectile dysfunction (IIEF-5 score <12) at twelve months was present among 74.4 % (baseline) and 75.1 % (ISPM) (p=0.90; Table 2) of patients, and incontinence (daily use of one or more urinary pads) was present among 27.6 % (baseline) and 27.7 % (ISPM) (p=0.98; Table 2).

## DISCUSSION

We found that implementing ISPM in a multidisciplinary tumour conference was associated with increased efficiency of the conference and less time needed to discuss each patient case. The results also indicated that the variability of the length of each case discussion was reduced with the use of ISPM. At the same time, the quality of the teamwork and the decision-making process was improved with the use of ISPM-MDT. These improvements were not, however, reflected in improved oncological or functional outcomes.

Using ISPM, discussing one patient took on average 72 seconds less, which corresponds to a 24 % reduction, or approximately 9-10 minutes shorter MDT in the current setting. Considering that there was a mean of 11.7 participants, more than one person-hour was saved during each session. This time saving is in agreement with results from another group developing a similar oncologic clinical decision support system for other cancer types.^4 9^

MDTs are in general scheduled events with a finite duration while the number of patient cases fluctuates and a structured process for the presentation and discussion of each case is paramount for retaining the quality of the decision making throughout the conference. We found both decreased variability of the duration of each case discussion and a consistent duration per case regardless of the number of patients discussed at the conference, indicating that the use of ISPM leads to a more structured and predictable process.

Although the quality of the MDTs increased when ISPM was used, this was not reflected in improved oncological or functional outcomes in our data. It should be noted that, already prior to the implementation of ISPM, the format of the preoperative prostate-cancer MDT conferences at Karolinska had been structuralized – although not into a digital format – with an apparent effect on the nerve-sparing strategy as well as on the risk of positive surgical margins.^10^

While it has been shown that MDTs lead to more accurate staging,^11^ higher adherence to clinical guidelines,^12^ and shorter time to treatment after diagnosis,^13^ several prior studies have failed to show improved outcome among patients discussed in MDT meetings^14-16^ while other have reported better outcomes.^17-19^ The MDT conference is a costly process, and it is important that future studies justify the costs through evidence of better outcomes.

A structured digital format for the MDT conference entails several potential further advantages apart from efficiency and quality in the decision making. For example, the resulting database can be used for real-time quality assessment, feedback to pathologists, radiologists, surgeons, and radiotherapists. Also, it enables development of prognostic models for better prediction tailored to the centers’ own patient cohorts. Patient-reported outcome measures can be used in the communication with the patient during follow up for a more structured care of the side effects of treatment and for spending more time with the patient on solving problems rather than understanding them. None of these advantages were assessed in the present study but are all strong potential benefits of a digital platform such as ISPM.

Without simultaneous evaluation of positive surgical margins and functional outcomes, quality assessment of prostate-cancer surgery is of little use since there is a reciprocal relation between radicality and postoperative function. Digital platforms connecting data points on all dimensions will facilitate more precise quality assessment. Ultimately, applying deep learning to make fuller use of these rich clinical, morphological, and patient-reported data is a promising future development.

The main limitation of this study is the observational design with non-concurrent comparison groups. The baseline measurement was carried out over of period of 33 months before the ISPM solution was implemented. Both treatment and outcome of prostate cancer change over time^20^ and differences between the baseline and ISPM periods may be attributable to other time-varying factors. For example, the lower frequency of pelvic lymph-node dissections in the ISPM period may, apart from a true effect of using the digital platform, be due to subtle changes in our operative indications for the procedure.

Access to patient-reported data is a major clinical need in healthcare in general, but particularly in the care of prostate cancer. The response rate to preoperative questionnaires was low in the baseline group, 32.6 % compared to 70.9 % the ISPM group. This difference reflects our effort made during the study period to increase patient participation in the national questionnaires on functional outcome rather than an effect of the digital platform.

## CONCLUSION

Our implementation of the ISPM clinical decision support system in MDT sessions at Karolinska University Hospital was associated with more efficient presentations and decision making in the conference as well as higher perceived quality of the decision process, but not with improved patient outcomes.

## Data Availability

All non-personal data produced in the present study are available upon reasonable request to the authors. All personal data used in the present study are withheld in accordance with Swedish legislation on patient data and GDPR.

## ACKNOWLEDGEMENTS

This project has received funding from the European Union’s Horizon 2020 research and innovation programme under grant agreement No 780495.

## REFERENCES

1. Pillay B, Wootten AC, Crowe H, et al. The impact of multidisciplinary team meetings on patient assessment, management and outcomes in oncology settings: A systematic review of the literature. Cancer Treat Rev 2016;42:56–72. doi: 10.1016/j.ctrv.2015.11.007 [published Online First: 2015/12/09]

2. Rosell L, Alexandersson N, Hagberg O, et al. Benefits, barriers and opinions on multidisciplinary team meetings: a survey in Swedish cancer care. BMC Health Serv Res 2018;18(1):249. doi: 10.1186/s12913-018-2990-4 [published Online First: 2018/04/07]

3. Farrugia DJ, Fischer TD, Delitto D, et al. Improved Breast Cancer Care Quality Metrics After Implementation of a Standardized Tumor Board Documentation Template. J Oncol Pract 2015;11(5):421–3. doi: 10.1200/JOP.2015.003988 [published Online First: 2015/09/19]

4. Hammer RD, Fowler D, Sheets LR, et al. A digital tumor board solution impacts case discussion time and postponement of cases in tumor boards. Health and Technology 2021;11(3):525–33. doi: 10.1007/s12553-021-00533-x

5. Brown GTF, Bekker HL, Young AL. Quality and Efficacy of Multidisciplinary Team (MDT) Quality Assessment Tools and Discussion Checklists: A Systematic Review. BMC Cancer, 2021.

6. Lamb BW, Wong HW, Vincent C, et al. Teamwork and team performance in multidisciplinary cancer teams: development and evaluation of an observational assessment tool. BMJ Qual Saf 2011;20(10):849–56. doi: 10.1136/bmjqs.2010.048660 [published Online First: 2011/05/26]

7. Rosen RC, Cappelleri JC, Smith MD, et al. Development and evaluation of an abridged, 5-item version of the International Index of Erectile Function (IIEF-5) as a diagnostic tool for erectile dysfunction. Int J Impot Res 1999;11(6):319–26. doi: 10.1038/sj.ijir.3900472 [published Online First: 2000/01/19]

8. Epstein JI, Egevad L, Amin MB, et al. The 2014 International Society of Urological Pathology (ISUP) Consensus Conference on Gleason Grading of Prostatic Carcinoma: Definition of Grading Patterns and Proposal for a New Grading System. Am J Surg Pathol 2016;40(2):244–52. doi: 10.1097/PAS.0000000000000530 [published Online First: 2015/10/23]

9. Krupinski EA, Comas M, Gallego LG, et al. A New Software Platform to Improve Multidisciplinary Tumor Board Workflows and User Satisfaction: A Pilot Study. J Pathol Inform 2018;9:26. doi: 10.4103/jpi.jpi_16_18 [published Online First: 2018/09/01]

10. Jaderling F, Akre O, Aly M, et al. Preoperative staging using magnetic resonance imaging and risk of positive surgical margins after prostate-cancer surgery. Prostate Cancer Prostatic Dis 2019;22(3):391–98. doi: 10.1038/s41391-018-0116-z [published Online First: 2018/12/07]

11. Sundi D, Cohen JE, Cole AP, et al. Establishment of a new prostate cancer multidisciplinary clinic: Format and initial experience. Prostate 2015;75(2):191–9. doi: 10.1002/pros.22904 [published Online First: 2014/10/14]

12. Korman H, Lanni T, Jr., Shah C, et al. Impact of a prostate multidisciplinary clinic program on patient treatment decisions and on adherence to NCCN guidelines: the William Beaumont Hospital experience. Am J Clin Oncol 2013;36(2):121–5. doi: 10.1097/COC.0b013e318243708f [published Online First: 2012/02/07]

13. Yopp AC, Mansour JC, Beg MS, et al. Establishment of a multidisciplinary hepatocellular carcinoma clinic is associated with improved clinical outcome. Ann Surg Oncol 2014;21(4):1287–95. doi: 10.1245/s10434-013-3413-8 [published Online First: 2013/12/10]

14. Boxer MM, Vinod SK, Shafiq J, et al. Do multidisciplinary team meetings make a difference in the management of lung cancer? Cancer 2011;117(22):5112–20. doi: 10.1002/cncr.26149 [published Online First: 2011/04/28]

15. Swellengrebel HA, Peters EG, Cats A, et al. Multidisciplinary discussion and management of rectal cancer: a population-based study. World J Surg 2011;35(9):2125–33. doi: 10.1007/s00268-011-1181-9 [published Online First: 2011/07/02]

16. Wille-Jorgensen P, Sparre P, Glenthoj A, et al. Result of the implementation of multidisciplinary teams in rectal cancer. Colorectal Dis 2013;15(4):410–3. doi: 10.1111/codi.12013 [published Online First: 2012/09/11]

17. Koco L, Weekenstroo HHA, Lambregts DMJ, et al. The Effects of Multidisciplinary Team Meetings on Clinical Practice for Colorectal, Lung, Prostate and Breast Cancer: A Systematic Review. Cancers (Basel) 2021;13(16) doi: 10.3390/cancers13164159 [published Online First: 2021/08/28]

18. MacDermid E, Hooton G, MacDonald M, et al. Improving patient survival with the colorectal cancer multi-disciplinary team. Colorectal Dis 2009;11(3):291–5. doi: 10.1111/j.1463-1318.2008.01580.x [published Online First: 2008/05/15]

19. Palmer G, Martling A, Cedermark B, et al. Preoperative tumour staging with multidisciplinary team assessment improves the outcome in locally advanced primary rectal cancer. Colorectal Dis 2011;13(12):1361–9. doi: 10.1111/j.1463-1318.2010.02460.x [published Online First: 2010/10/21]

20. Orrason AW, Westerberg M, Garmo H, et al. Changes in treatment and mortality in men with locally advanced prostate cancer between 2000 and 2016: a nationwide, population-based study in Sweden. Bju Int 2020;126(1):142–51. doi: 10.1111/bju.15077 [published Online First: 2020/04/11]

