## Supplementary Table 1 for "The effect of digital-enabled multidisciplinary therapy conferences on efficiency and quality of the decision making in prostate-cancer care"

**Supplementary Table 1:** Modified MDT-MODe version used in this MDT study

| **MDT-MODe Response Options** | **A** | **B** | **C** |
| --- | --- | --- | --- |
| **Patient's view** | No knowledge of patient's wishes or opinions regarding treatment | Vague first-hand knowledge, or good second-hand knowledge of patient's wishes or opinions regarding treatment | Comprehensive first-hand knowledge of patient's wishes or opinions regarding treatment |
| **Psychosocial** | No knowledge of patient's personal circumstances, social and psychological issues | Vague first-hand knowledge, or good second-hand knowledge of patient's personal circumstances, social and psychological issues | Comprehensive first-hand knowledge of patient's personal circumstances, social and psychological issues |
| **Co-morbidity** | No knowledge of past medical history and performance status | Vague first-hand knowledge, or good second-hand knowledge of past medical history and performance status | Comprehensive first-hand knowledge of past medical history and performance status |
| **Pathology** | No provision of histopathological information | Histopathological information from a report / account | Histopathological information from pathologist |
| **Imaging** | No provision of radiological information | Radiological information from a report / account | Radiological images from radiologist |
| **History** | No patient case history | Partial case history | Fluent, comprehensive case history |
| **Decision** | No decision / decision unclear | Decision to defer to next MDT or partially clear decision | Clear decision about treatment(s) to be offered |
| **Members** | No contribution | Contribution inarticulate of vague | Clear contribution of specialty |
| **Chair** | Poor/inadequate leadership impeded team discussion and decision-making | Leadership neither enhanced or impeded team discussion and decision-making | Good leadership enhanced team discussion and decision-making |
| **MDT-MODe scores per response option** | **1** | **3** | **5** |
| **Participation in discussion** | Proportion of staff members participating in the discussion [%] | | |
