## Supplementary Table 2 for "The effect of digital-enabled multidisciplinary therapy conferences on efficiency and quality of the decision making in prostate-cancer care"

**Supplementary Table 2:** Full description of oncological and perioperative outcomes

|  | **Baseline** | **ISPM** | **p-value** |
| --- | --- | --- | --- |
| **Positive surgical margin** | | | LRT 0.51 |
| Frequencies | 251/906 (27.7 %) [#missing^1^ = 18] | 98/378 (25.9 %) [#missing^1^ = 27] |  |
| Prevalence ratios | 1.0 (Ref.) | 0.94 (0.76 - 1.14) |  |
| **Significant positive margin status (among positive margins)** | | | LRT 0.94 |
| Frequencies | 101/251 (40.2 %) [#missing = 0] | 39/98 (39.8 %) [#missing = 0] |  |
| Prevalence ratios | 1.0 (Ref.) | 0.99 (0.73 - 1.30) |  |
| **Significant positive margin status (entire population)** | | | LRT 0.66 |
| Frequencies | 101/906 (11.1 %) [#missing^1^ = 18] | 39/378 (10.3 %) [#missing^1^ = 27] |  |
| Prevalence ratios | 1.0 (Ref.) | 0.93 (0.64 - 1.30) |  |
| **Nerve-sparing right** | | | M-W 0.36 |
| intra | 286 (31.3 %) | 114 (29.5 %) |  |
| inter | 343 (37.6 %) | 145 (37.5 %) |  |
| semi | 127 (13.9 %) | 52 (13.4 %) |  |
| no nerve-sparing - right | 157 (17.2 %) | 76 (19.6 %) |  |
| missing^1^ | 11 (1.2 %) | 18 (4.4 %) |  |
| **Nerve-sparing right - dichotomized - outcome: no nerve-sparing** | | | LRT 0.49 |
| Frequencies | 284/913 (31.1 %)  [#missing^1^ = 11 (1.2 %)] | 128/387 (33.1 %)  [#missing^1^ = 18 (4.4 %)] |  |
| Prevalence ratios | 1.0 (Ref.) | 1.06 (0.89 - 1.26) |  |
| **Nerve-sparing left** | | | M-W 0.29 |
| intra | 271 (29.7 %) | 106 (27.4 %) |  |
| inter | 297 (32.5 %) | 128 (33.1 %) |  |
| semi | 159 (17.4 %) | 61 (15.8 %) |  |
| no nerve-sparing - left | 186 (20.4 %) | 92 (23.8 %) |  |
| missing | 11 (1.2 %) | 18 (4.4 %) |  |
| **Nerve-sparing left - dichotomized - outcome: no nerve-sparing** | | | LRT 0.55 |
| Frequencies | 345/913 (37.8 %)  [#missing^1^ = 11 (1.2 %)] | 153/387 (39.5 %)  [#missing^1^ = 18 (4.4 %)] |  |
| Prevalence ratios | 1.0 (Ref.) | 1.05 (0.90 - 1.21) |  |
| **Nerve-sparing left or right - compilation of dichotomized versions - with pairwise comparisons** | | | χ^2^ = 0.005 - OVERALL |
| bilateral | 392 (42.9 %) | 140 (36.2 %) | χ^2^ = 0.05^2^ |
| unilateral | 413 (45.2 %) | 213 (55.0 %) | χ^2^ = 0.005^2^ |
| no nerve-sparing | 108 (11.8 %) | 34 (8.8 %) | χ^2^ = 0.13^2^ |
| missing^1^ | 11 (1.2 %) | 18 (4.4 %) |  |
| **Nerve-sparing - any** | | | M-W = 0.11 |
| any nerve-sparing | 805 (88.2 %) | 353 (91.2 %) |  |
| no nerve-sparing | 108 (11.8 %) | 34 (8.8 %) |  |
| missing^1^ | 11 (1.2 %) | 18 (4.4 %) |  |
| **Pelvic lymph-node dissection** | | | M-W p < 0.001 |
| no | 543 (60.0 %) | 248 (70.1 %) |  |
| yes | 362 (40.0 %) | 106 (29.9 %) |  |
| missing^1^ | 19 (2.1 %) | 51 (12.6 %) |  |
| **Lymph-node metastases among patients that underwent pelvic lymph-node dissection** | | | M-W p = 0.92 |
| no (N0) | 302 (83.4 %) | 88 (83.0 %) |  |
| yes (N1) | 60 (16.6 %) | 18 (17.0 %) |  |
| missing^1^ | 0 (0 %) | 0 (0 %) |  |
| *1 – percentage missing calculated on the entire cohort. 2 – pairwise comparison; Bonferroni-Holm corrected for multiple testing* | | | |
