## Supplementary Table 3 for "The effect of digital-enabled multidisciplinary therapy conferences on efficiency and quality of the decision making in prostate-cancer care"

**Supplementary Table 3:** Full description of functional (urinary, sexual) outcomes

|  | **Baseline** | **ISPM** | **p-value** |
| --- | --- | --- | --- |
| **Incontinence at 12 months post-surgery among all individuals** | | | LRT 0.98 |
| Frequencies | 171/619 (27.6 %) [#missing^1^ = 305 (33 %)] | 64/231 (27.7 %) [#missing^1^ = 174 (43 %)] |  |
| Prevalence ratios | 1.0 (Ref.) | 1.0 (0.78 - 1.27) |  |
| **Incontinence at 12 months among individuals who reported being continent preoperatively** | | | LRT 0.94 |
| Frequencies | 62/218 (28.4 %) [#missing^1^ = 78 (26.4 %)] | 51/177 (28.8 %) [#missing^1^ = 112 (38.8 %)] |  |
| Prevalence ratios | 1.0 (Ref.) | 1.01 (0.74 - 1.38) |  |
| **Postoperative IIEF-5 score at 12 months among all individuals** | | | M-W 0.35 |
|  | mean = 7.5 (SD = 7.8)  [#missing^1^ = 320 (34.6 %)] | mean = 7.2 (SD = 7.7)  [#missing^1^ = 176 (43.5 %)] |  |
| **Erectile dysfunction at 12 months among all individuals - outcome: impotence (IIEF-5 score <12)** | | | LRT 0.90 |
| Frequencies | 451/604 (74.7 %) [#missing^1^ = 320 (34.6 %)] | 172/229 (75.1 %) [#missing^1^ = 176 (43.5 %)] |  |
| Prevalence ratios | 1.0 (Ref.) | 1.01 (0.92 - 1.09) |  |
| **Erectile dysfunction at 12 months among individuals who reported being potent preoperatively - outcome: impotence** | | | LRT 0.34 |
| Frequencies | 70/126 (55.6 %) [#missing^1^ = 47 (27.2 %)] | 63/102 (61.8 %) [#missing^1^ = 60 (37.0 %)] |  |
| Prevalence ratios | 1.0 (Ref.) | 1.11 (0.89 - 1.39) |  |
| **Urinary function bother at 12 months** | | | M-W 0.49 |
| Not at all | 285 (46.3) | 113 (48.9) |  |
| A little | 212 (34.4) | 78 (33.8) |  |
| Moderately | 73 (11.9) | 20 (8.7) |  |
| A lot | 46 (7.5) | 20 (8.7) |  |
| Missing^1^ | 308 (33.3 %) | 174 (43 %) |  |
| **Urinary function bother at 12 months - dichotomized - outcome: no bother or a little bother** | | | LRT 0.50 |
| Frequencies | 497/616 (80.7 %) [#missing^1^ = 308 (33.3 %)] | 191/231 (82.7 %) [#missing^1^ = 174 (43 %)] |  |
| Prevalence ratios | 1.0 (Ref.) | 1.02 (0.95 - 1.10) |  |
| **Erectile function bother at 12 months** | | | M-W 0.77 |
| Not at all | 98 (15.9) | 38 (16.6) |  |
| A little | 179 (29.0) | 62 (27.1) |  |
| Moderately | 175 (28.4) | 64 (27.9) |  |
| A lot | 165 (26.7) | 65 (28.4) |  |
| Missing^1^ | 307 (33.2 %) | 174 (43 %) |  |
| **Erectile function bother at 12 months - dichotomized - outcome: no bother or a little bother** | | | LRT 0.75 |
| Frequencies | 277/617 (44.9 %) [#missing^1^ = 307 (33.2 %)] | 100/229 (43.7 %) [#missing^1^ = 176 (43.5 %)] |  |
| Prevalence ratios | 1.0 (Ref.) | 0.97 (0.81 - 1.15) |  |

*1 – percentage missing calculated on the entire cohort.*
